# Defining Mechanistic Links Between the Non-Coding Variant rs17673553 in *CLEC16A* and Lupus Susceptibility

**DOI:** 10.1101/2024.12.02.24318337

**Authors:** Harikrishna Reddy-Rallabandi, Manish K. Singh, Loren L. Looger, Swapan K. Nath

## Abstract

Systemic lupus erythematosus (SLE) is a complex autoimmune disorder characterized by widespread inflammation and autoantibody production. Its development and progression involve genetic, epigenetic, and environmental factors. Although genome-wide association studies (GWAS) have repeatedly identified a susceptibility signal at 16p13, its fine-scale source and its functional and mechanistic role in SLE remain unclear. We used bioinformatics to prioritize likely functional variants and validated the top candidate through various experimental techniques, including CRISPR-based genome editing in B cells. To assess the functional impact of the proposed causal variant in *CLEC16A*, we compared autophagy levels between wild-type (WT) and knock-out (KO) cells. Systematic bioinformatics analysis identified the highly conserved non-coding intronic variant rs17673553, with the risk allele apparently affecting enhancer function and regulating several target genes, including *CLEC16A* itself. Luciferase reporter assays followed by ChIP-qPCR validated this enhancer activity, demonstrating that the risk allele increases the binding of enhancer histone marks (H3K27ac and H3K4me1), CTCF-binding factor, and key immune transcription factors (GATA3 and STAT3). Knock-down of *GATA3* and *STAT3 via* siRNA led to a significant decrease in *CLEC16A* expression. These regulatory effects on the target gene were further confirmed using CRISPR-based genome editing and CRISPR-dCas9-based epigenetic activation/silencing. Functionally, WT cells exhibited higher levels of starvation-induced autophagy compared to KO cells, highlighting the role of *CLEC16A* and the rs17673553 locus in autophagy regulation. These findings suggest that the rs17673553 locus – particularly the risk allele – drives significant allele-specific chromatin modifications and binding of multiple transcription factors, thereby mechanistically regulating the expression of target autophagy-associated genes, including *CLEC16A* itself. This mechanism could potentially explain the association between rs17673553 and SLE, and underlie the signal at 16p13.

## Introduction

Systemic lupus erythematosus (SLE) is a complex autoimmune disorder characterized by chronic inflammation and the production of autoantibodies against the body’s own tissues[1]. This multisystem disease is highly heterogeneous, presenting a broad spectrum of clinical manifestations ranging from mild, self-resolving symptoms to severe, life-threatening multi-organ damage. This variability in disease presentation contributes to significant morbidity and complicates the clinical diagnosis and management of SLE.

The etiology of SLE is multifactorial, resulting from a dynamic interplay between genetic predisposition, epigenetic changes, and environmental triggers[2,3]. Although considerable progress has been made in identifying genetic variants associated with SLE, understanding the precise molecular mechanisms that drive disease development and progression remains a significant challenge. Numerous genes contribute to immune dysregulation in SLE. These genetic factors are further modulated by epigenetic alterations, such as DNA methylation and histone modifications.

Despite advances in our understanding of SLE, many aspects of its pathogenesis remain poorly understood. Ongoing research efforts are focused on unraveling the molecular pathways involved in disease development and progression, with the ultimate goals of identifying novel therapeutic targets, understanding diverse subclinical manifestations, and improving patient outcomes.

Multiple genome-wide association studies (GWAS) have identified numerous loci associated with SLE susceptibility, including the 16p13 locus[4]. Initially discovered in 2007 as a type 1 diabetes (T1D) susceptibility peak[5], this locus has since been linked to various autoimmune diseases, including multiple sclerosis (MS), rheumatoid arthritis (RA), and SLE. The broad association with autoimmunity suggests that this locus may contribute to common pathogenic pathways underlying autoimmune disorders. It was first reported as an SLE risk locus in 2009 in a Chinese population[6], and since then, several studies, including ours, have reported broad genetic association with SLE across Asian, European, and Hispanic ancestries. Despite its consistent identification in genetic studies, the functional and mechanistic roles of this locus in SLE pathogenesis remain poorly understood. It is hypothesized that variants within this region may influence the expression of key immune regulatory genes, although the precise pathways and regulatory networks affected by these variants are yet to be clarified[7-9].

Among the genes at this locus, C-type lectin domain family 16, member A (*CLEC16A*) is the most interesting. As the name indicates, CLEC16A was originally annotated as a C-type lectin; however, it has since been discovered to instead function as an endolysosomal E3 ubiquitin ligase that regulates pancreatic β-cell mitophagy, among other roles[10]. *CLEC16A* and its polymorphisms have been implicated in several autoimmune diseases, including common variable immunodeficiency disorder (CVID)[11], RA and juvenile idiopathic arthritis[12], MS[13], and others. Although the exact biological roles of *CLEC16A* are still being elucidated, it is known to be involved in many critical cellular processes, including autophagy, mitophagy, endocytosis, intracellular trafficking, and immune system regulation[14,15]. Dysregulation of autophagy – a process vital for maintaining cellular homeostasis and regulating immune responses – has been linked to various autoimmune conditions, including SLE[16-18]. However, the specific roles of *CLEC16A* and its genetic variants in autophagy regulation and SLE pathogenesis remain largely unexplored.

In this study, we focused on a highly conserved non-coding intronic variant, rs17673553, which has been identified as a potential risk allele for SLE. By employing bioinformatics prioritization, genome editing techniques, and cellular assays, we sought to elucidate how this enhancer variant affects *CLEC16A* expression and autophagy regulation in B-cells. We aimed to uncover the molecular mechanisms linking rs17673553 to *CLEC16A* expression level and protein activity and autophagy, shedding light on the functional significance of this genetic variant and its role in the pathogenesis of SLE.

## Results

### Identification of rs17673553 as a functional variant at the 16q13 locus

In our ImmunoChip-based SLE association study[19] across multiple Asian populations (Korean, Chinese, and Malaysian), we identified 10 novel loci and replicated several known loci (including the 16p13 locus) at genome-wide significance (p<5×10^−^□). At 16p13, we reported the strongest association with rs35032408, a SNP located within intron 23 of *CLEC16A*. This discovery prompted us to focus on identifying and validating potentially functional SNPs within the haplotype block containing rs35032408.

rs35032408 exhibits weak linkage disequilibrium (LD) with previously reported SLE-associated SNPs (**Supplementary Table ST1, Supplementary Figure 1A**). Using data from the 1000 Genomes Project, we identified six common SNPs within a strong LD block spanning ∼28.8 kb. Through comprehensive bioinformatics analysis, we explored potential regulatory elements within this target LD region, utilizing publicly available datasets from ENCODE and other databases to identify active enhancers and regulatory regions **(Supplementary Figure 1B)**.

We next evaluated the functional potential of each corrected SNP using predicted deleteriousness scores. Two SNPs, rs17673553 and rs62026379, were predicted to be deleterious using PredictSNP2 (**Supplementary Table ST1)**. Notably, rs17673553 is highly conserved and has a high deleteriousness score (17.4) in CADD (Combined Annotation-Dependent Depletion)[20], indicating significant pathogenic potential. To refine our assessment of causality, we utilized PICS2 (Probabilistic Identification of Causal SNPs), which estimated a higher causality for rs17673553 (0.32) than the nearest LD SNP, rs62026379 (0.12) (**Supplementary Table ST2**). Based on these analyses, rs17673553 was selected as the top candidate for experimental follow-up.

Analysis using GTEx and eQTLgen identified rs17673553 as an expression quantitative trait locus (eQTL) for multiple genes, including *CLEC16A*. Additionally, we annotated regions topologically associated with rs17673553 using ENCODE Hi-C/Micro-C chromatin-conformation data (**Supplementary Figure 2**), which showed that the rs17673553 region associates with the *CLEC16A* promoter and overlaps with the *CIITA* (Class II Major Histocompatibility Complex Transactivator) gene.

### Impact of rs17673553 on enhancer activity

We evaluated allele-specific enhancer activity of a 281-bp region surrounding rs17673553 (containing either the risk or non-risk SNP) using dual-luciferase reporter assays in both non-immune HEK293 (embryonic kidney) cells and immune cells, including Namalwa (B-lymphocyte) and Jurkat (T-lymphocyte) cells. I all cell types tested, the rs17673553 risk allele (AA) showed significantly higher enhancer activity than the non-risk allele (GG) (p-values: <0.0001, <0.001, 0.016, respectively; **Figure 1A**). These results demonstrate that the rs17673553 risk allele significantly increases enhancer activity across different cell types, underscoring the functional relevance of this SNP.

**Figure 1.**
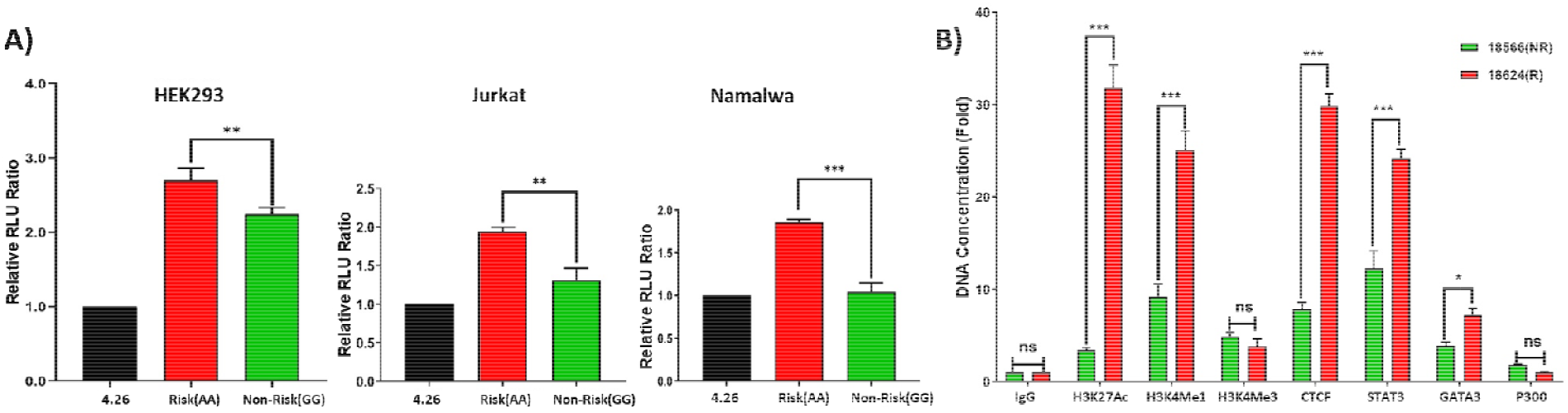
Allele-specific activity of rs17673553. **A)** Luciferase reporter assay results show allele-specific enhancer activity in non-immune (HEK293) and immune (Namalwa, Jurkat) cells. The risk allele (AA) showed significantly higher (Namalwa, p<0.0001; Jurkat, p<0.001; and HEK293, p<0.01) expression than the non-risk (GG) allele. **B)** ChIP assay results showing allele-specific binding to DNA-binding proteins. Histone marks H3K27ac (p<0.0001) and H3K4me1 (p<0.0001), CTCF (p<0.0001), STAT3 (p<0.0001) and GATA3 (p<0.01) showed significantly higher binding to risk allele (AA) than non-risk allele (GG). Data represent mean ± SD from three biological replicates (n = 3). Statistical analysis was performed using GraphPad Prism.

Using ChIP-grade antibodies, we then investigated the binding patterns of three regulatory histone marks— H3K27ac, H3K4me1, and H3K4me3—at the SNP locus within its biological context. Binding quantification was performed using ChIP-qPCR (**Figure 1B**). We observed significantly higher binding levels for two enhancer marks in GM18624 (risk AA genotype) compared to GM18566 (non-risk GG genotype): H3K27ac showed a ∼9-fold increase (p<0.0001) and H3K4me1 showed a ∼3-fold increase between risk and non-risk (p<0.0001). In contrast, H3K4me3 (a promoter mark) showed no significant or allele-specific differences. Additionally, we tested several DNA-binding proteins and transcription factors (TFs) suggested by bioinformatics (**Supplementary Figure 4A, B**). CTCF, GATA3, and STAT3 showed significant binding differences, binding more to GG non-risk: CTCF >3-fold (p<0.0001), STAT3 >1.5-fold (p<0.0001) and GATA3 >1.3-fold (p<0.01) (**Figure 1B**).

These findings provide strong evidence that rs17673553 is located within an active enhancer region, where interactions among RNA polymerase, histone modifications, chromatin regulators, and allele-specific elements collectively influence transcriptional regulation – with the risk allele recruiting dramatically more transcriptional activators and thus likely leading to higher expression of *CLEC16A* and other downstream target genes.

### CRISPR-based epigenetic modification around rs17673553 modulates *CLEC16A* expression

Epigenetic modifications in regulatory regions play crucial roles in regulating cognate target gene expression. Above we identified dramatic changes in epigenetic marks around the risk and non-risk rs17673553 alleles. To evaluate whether such epigenetic modifications around rs17673553 could modulate target genes including *CLEC16A* itself, we used CRISPR-based inhibition and activation (CRISPRa/i)[21]. Specifically, we employed CRISPR/dCas9-VPR and dCas9-TET1 to activate[22], and dCas9-DNMT3A and dCas9-MeCP2 to inhibit[21], transcriptional activity around rs17673553. VPR and TET1 activation strongly increased (∼1.5- and 1.8-fold, p<0.0001) *CLEC16A* expression in Namalwa cells (**Figure 2**), while DNMT3A and MECP2-mediated inhibition reduced *CLEC16A* expression by ∼30% (p<0.0001; **Figure 2**). These findings provide mechanistic insights into how rs17673553 affects the expression of *CLEC16A* and neighboring genes.

**Figure 2.**
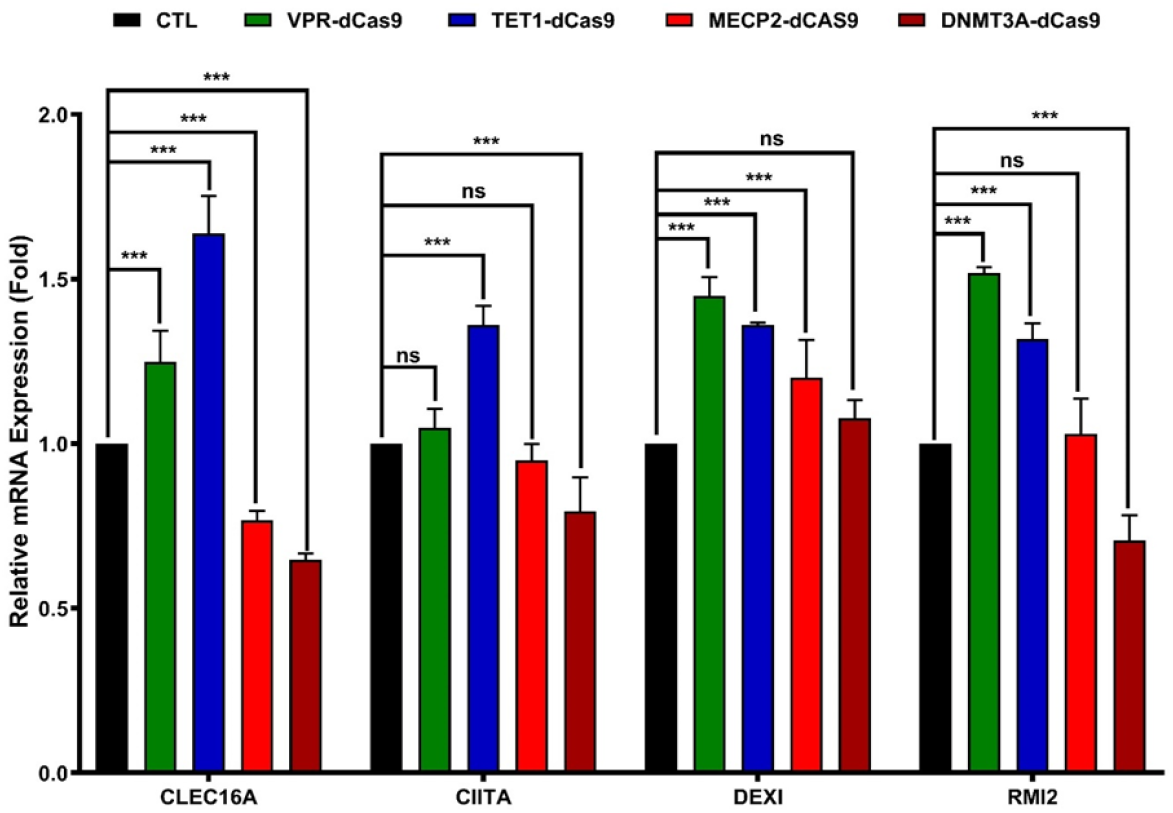
CRISPR-based epigenetic modification. CRISPR-dCas9 plasmids were co-transfected along with the guide RNA cloned vector. qRT-PCR results show that activation dCas9-vectors VPR significantly enhanced the expression of *CLEC16A* (p<0.0001), *DEXI* (p <0.0001) and *RMI2* (p<0.0001); no notable impact was observed on *CIITA* (p=0.63). TET1 significantly increased the expression of *CLEC16A* (p<0.0001), *CIITA* (p<0.0001), *DEXI* (p<0.0001), and *RMI2* (p<0.0001). Inhibitory dCas9-vectors MECP2 significantly suppressed *CLEC16A* (p<0.0001) and *DEXI* (p<0.0001), with no observable effects on *CIITA* (p=0.58) and *RMI2* (p=0.89). Similarly, DNMT3A suppressed *CLEC16A* (p<0.0001), CIITA (p<0.0001), and *RMI2* (p<0.0001) expression, with no significant impact on *DEXI* (p=0.23). Data represent mean ± SD from three biological replicates (n = 3) and analyzed using two-way ANOVA in GraphPad Prism.

These results demonstrate that the rs17673553 locus profoundly impacts *CLEC16A* expression in Namalwa cells. Effects on nearby genes (*CIITA*; *DEXI*, dexamethasone-induced protein 1 – a transcriptional regulator of poorly understood function; *RMI2*, RecQ-mediated genome instability 2 topoisomerase subunit) were mixed. Transcriptional activators significantly increased expression of nearby genes, as expected. Transcriptional repressors lessened expression of *CIITA* and *RMI2* but increased levels of *DEXI*, suggesting a complex interplay of the regulatory elements in this region. Overall, the strongest effects – both for activation and inhibition – were on *CLEC16A* itself.

### Validating transcriptional effects of the rs17673553 locus using CRISPR/Cas9

To directly assess the regulatory impact of the rs17673553 locus, we employed CRISPR/Cas9 using a 2- sgRNA pool to delete 183 base pairs around this SNP in Namalwa cells (**Methods**). After serial dilution and selection, we generated two isogenic lines: uncut wild-type (WT) and rs17673553 knockout (KO) (**Figure 3A, Supplementary Figure 3**).

**Figure 3.**
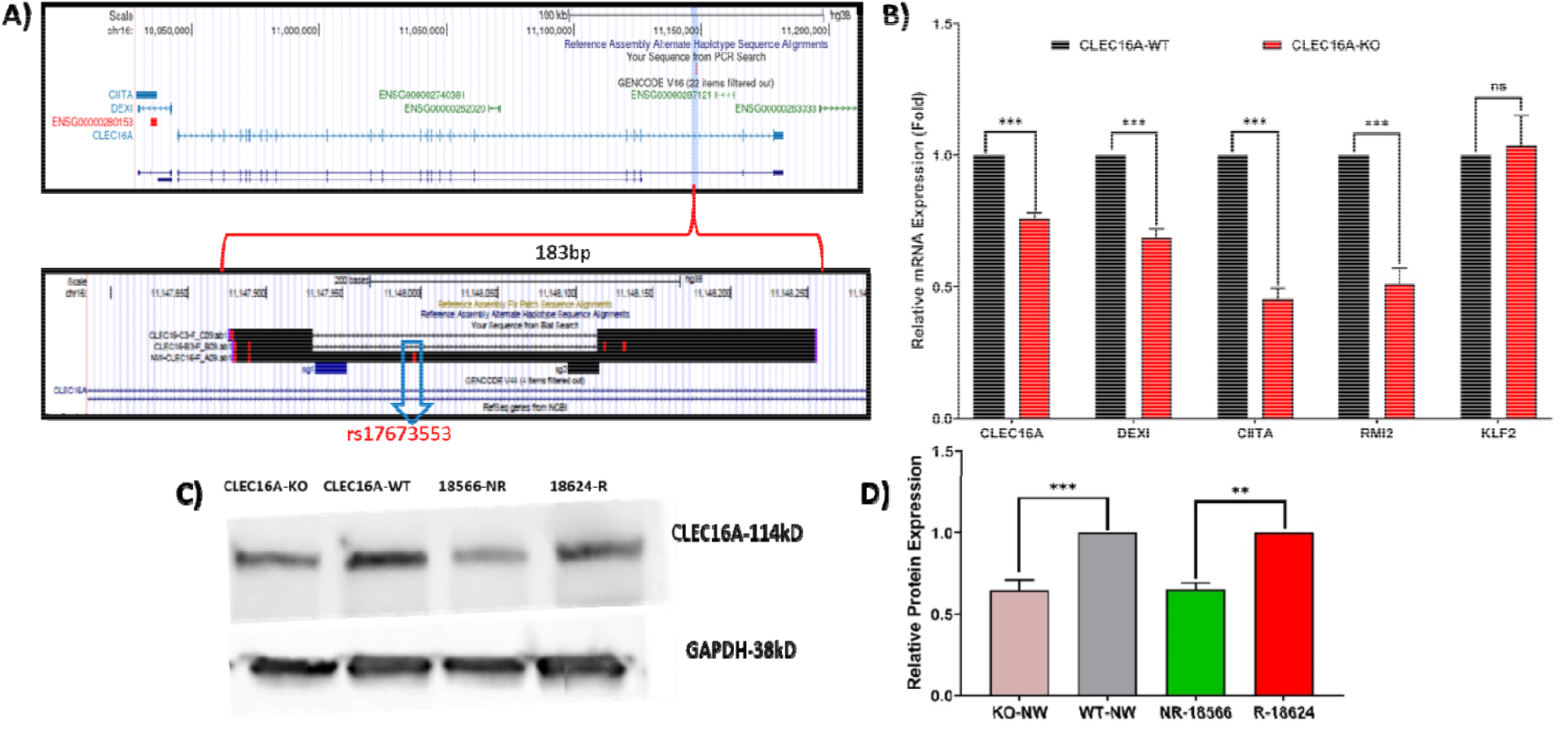
CRISPR knock-out of *CLEC16A*. **A)** A 183bp sequence surrounding rs17673553 was knocked-out using CRISPR-Cas9. Genomic coordinates of CLEC16A (upper panel) and the intronic SNP. Comparative results (lower panel) showing the 2 sgRNAs and the CRSIPR-knockout region from CLEC16A-KO cells and SNP- containing sequence from wild-type cells. **B)** qRT-PCR results showing the effect of the SNP region knockout on target gene *CLEC16A* and other nearby genes. Significantly lower expression of target gene *CLEC16A* (p<0.0001) and adjacent genes *DEXI* (p<0.0001), *CIITA* (p<0.001) and *RMI2* (p<0.0001) was seen in KO cells than WT cells. A neighboring control gene *KLF2* (p=0.999) had no significant impact on its expression. **C)** Western blot analysis showing the differential expression of *CLEC16A* in KO (lane1) and WT (lane2) cells, as well as in non-risk (18566) and risk (18624) LCL cells. **D)** Densitometry analysis of Western blotting showing CLEC16A protein expression difference (fold) between WT-KO and risk/non-risk samples relative to a GAPDH control. Data represent mean ± SD from at least three biological replicates (n = 3).

We then measured target gene expression in WT and KO cells using qPCR. Compared to WT, target gene expression in KO cells was significantly decreased (**Figure 3B**). The expression level of *CLEC16A* was reduced by 30% (p<0.01), and neighboring genes all also decreased: *DEXI1* by 30% (p<0.001); *RMI2* by 50%, (p<0.0001); and *CITA* by 50% (p<0.001). As expected, the expression of the negative-control neighboring gene *KLF2* was unaffected. Thus, the rs17673553 locus helps govern expression of *CLEC16A, DEXI1, RMI2*, and *CITA*, but not of control genes.

In support of the qPCR results, quantitative Western blot (WB) analysis showed higher levels of CLEC16A protein in WT cells than KO cells (**Figure 3C**). When we compared the expression levels between two Han Chinese lymphoblastoid cell lines (Coriell NA18624 and NA18566) with risk (R) and non-risk (NR) genotype, respectively, we also found significant allele-specific differences, with the R allele associated with higher expression levels (**Figure 3D**) – consistent with qPCR results (**Figure 1A**). These findings demonstrate that the rs17673553 region significantly influences *CLEC16A* transcript and protein levels, and that it does so in an allele-dependent fashion, with the risk allele associated with higher transcript and protein levels of CLEC16A.

### siRNA-based knockdown of GATA3 and STAT3 reduces CLEC16A expression

Experimental ChIP-qPCR analysis and bioinformatic prediction identified GATA3 (**Figure 4**) and STAT3 (**Figure 5**) as key transcription factors binding near rs17673553 (**Supplementary Figure 4A, B**). Given this, we conducted siRNA-based knockdown experiments on these transcription factors in Namalwa cells to directly assess their effects on *CLEC16A* expression. Following transfection of siRNA against either factor, qPCR analysis showed a sharp and significant reduction in *CLEC16A* mRNA levels (**Figures 4, 5**). Specifically, *GATA3* knockdown led to a ∼40% decrease, and *STAT3* knockdown resulted in a ∼35% decrease, in *CLEC16A* expression compared to control cells treated with off-target siRNA. These findings suggest that both GATA3 and STAT3 positively regulate *CLEC16A* expression, likely through direct binding to enhancer elements near the rs17673553 locus.

**Figure 4.**
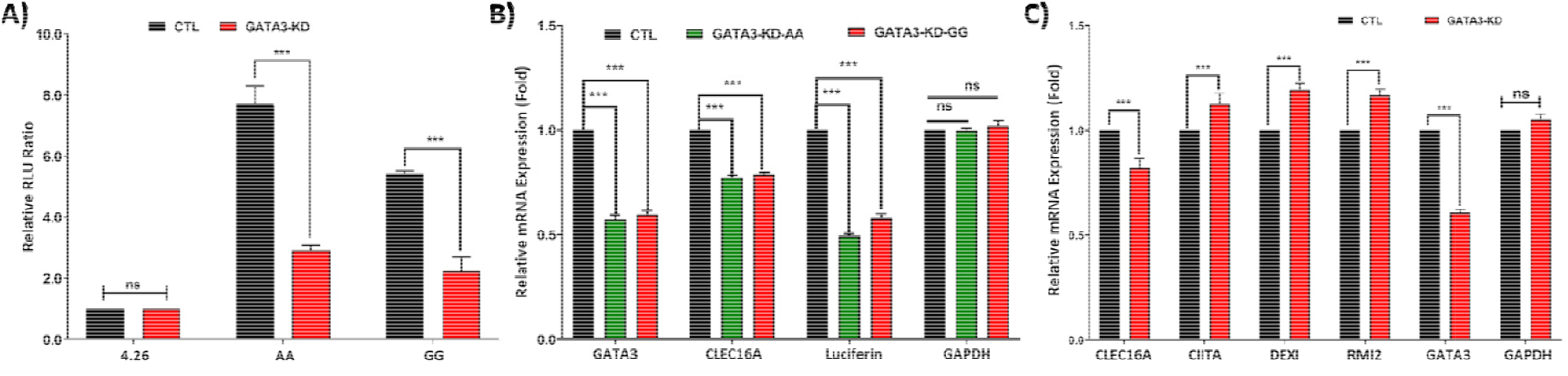
siRNA-based *GATA3* knock-down. To investigate the role of GATA3 at the rs17673553 locus, GATA3 was knocked down using siRNA in Namalwa cells. **A)** A combined experiment of siRNA-based knockdown and luciferase reporter assay results showing a significant decline in luciferase expressio risk-AA (p<0.0001) and non-risk-GG (p<0.0001) compared to control. **B)** qR-PCR of *luciferase*, GATA3 in both *GATA3, CLEC16A* and *GAPDH*. siRNA transfection significantly reduced *GATA3* (p<0.0001, <0.0001), *CLEC16A* (p<0.0001,0.0001) and *luciferase* (p<0.0001, <0.0001), with no difference observed in negative control *GAPDH*. **C)** qRT-PCR analysis of *GATA3* knockdown in Namalwa cells showed significant reduction in *CLEC16A* (p<0.0001) and *GATA3* (p<0.0001), and an increase in nearby genes *CIITA* (p<0.0001), *DEXI* (p<0.0001) and *RMI2* (p<0.0001), with housekeeping gene *GAPDH* showing no difference. Data represent mean ± SD, values and statistical significance calculated using two-way ANOVA in GraphPad Prism.

**Figure 5.**
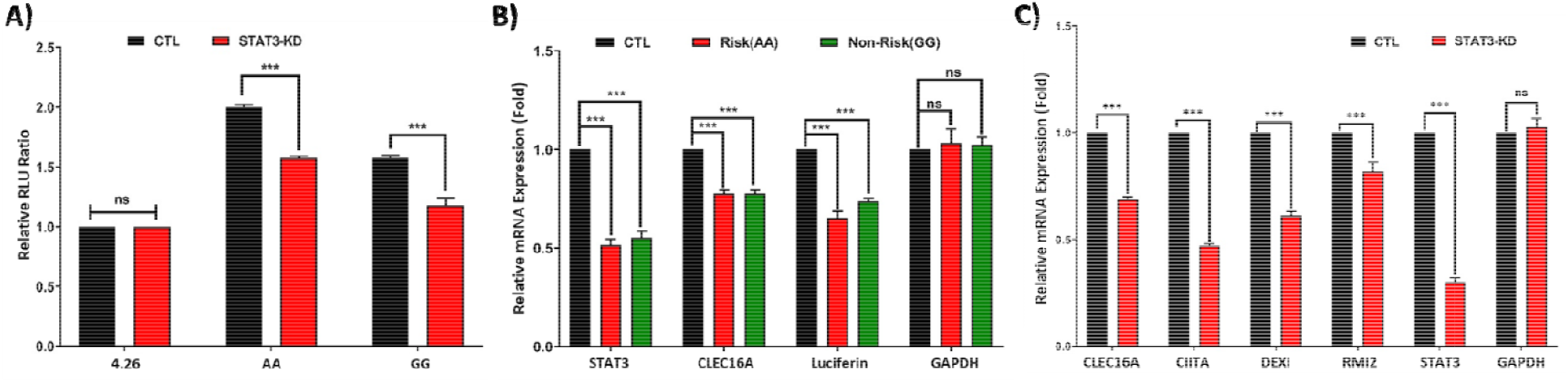
siRNA-based STAT3-Knockdown. To investigate the role of STAT3 at the rs17673553 locus, STAT3 was knocked down using siRNA in Namalwa cells. **A)** A combined experiment of siRNA-based STAT3 knockdown and luciferase reporter assay results showing a significant decline in luciferase expression in both risk-AA (p<0.0001) and non-risk-GG (p<0.0001) compared to control. **B)** qR-PCR of *luciferase, GATA3, CLEC16A* and *GAPDH*. siRNA transfection significantly reduced *STAT3* (p<0.0001, <0.0001), *CLEC16A* (p<0.0001,0.0001) and *luciferase* (p<0.0001, <0.0001), with no difference observed in negative control *GAPDH*. **C)** Comparative WT and STAT3-KD qRT-PCR results of *STAT3* knockdown in Namalwa cells showing strong reduction in the genes surrounding rs17673553 including *CLEC16A* (p<0.0001), *CIITA* (p<0.0001), *DEXI*(p<0.0001) and *RMI2*(p<0.0001) along with *STAT3*(p<0.0001), with housekeeping gene *GAPDH* showing no difference. Data represent mean ± SD from at least three biological replicates (n = 3), values and statistical significance calculated using two-way ANOVA in GraphPad Prism.

### CLEC16A negatively regulates starvation-induced autophagy by interacting with mTORC1

Given the established roles of CLEC16A in autophagy[14,15], we sought to establish the contribution of activity at the rs17673553 locus in mediating this activity. As such, we tested our WT and KO cells for starvation- induced autophagy by growth in Earle’s Balanced Salt Solution (EBSS) for six hours (**Figure 6A**). Quantification of several autophagic markers by qRT-PCR showed that KO cells showed significant upregulation over WT cells of some autophagy-related genes, including microtubule-associated proteins 1A/1B light chain 3A and 3B (*LC3A*, ∼1.5-fold, p<0.0001; *LC3B*, ∼1.5-fold, p<0.0001) and Unc-51-like autophagy- activating kinase 1 (*ULK1*, 2.2-fold, p<0.0001). Conversely, other autophagy-related genes, including sequestosome-1 (*SQSTM1, a*.*k*.*a. P62*, 0.25-fold, p<0.0001) and *CLEC16A* (0.5-fold, p< 0.0001), were significantly downregulated in KO cells.

**Figure 6A.**
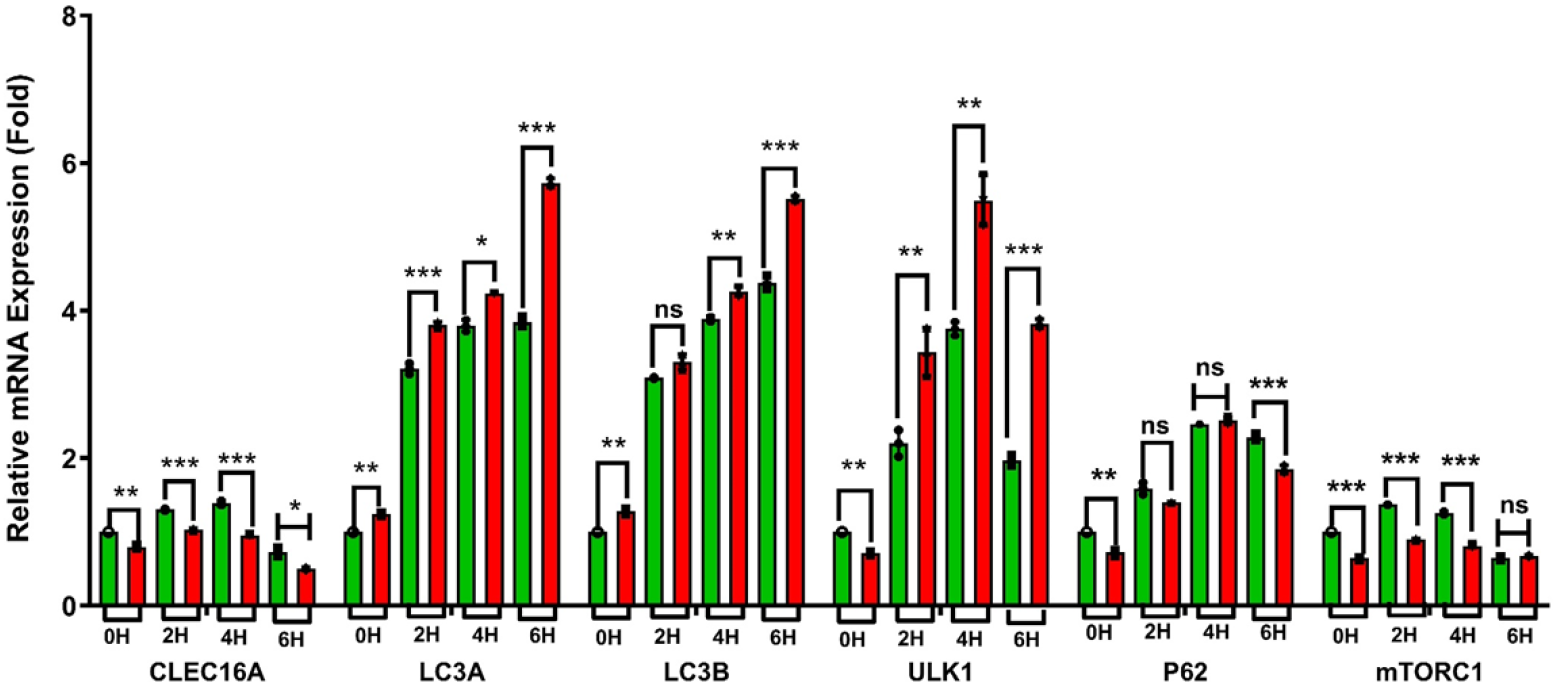
Starvation-induced autophagy. qRT-PCR assessment of EBSS-starved CLEC16A-WT and CLEC16-KO Namalwa cells showing differential expression of autophagy-related genes. Both WT and *CLEC16A- KO* cells were starved and analyzed for the samples at 0 hours (basal), 2hours, 4hours and 6hour regular intervals. Pairwise comparison between WT and KO showing *CLEC16A* expression is reduced across the samples at 0Hr slightly low (p=0.003), significant reduction at 2Hr (p<0.001), 4Hr (p<0.001) and at 6Hr it is greatly reduced in both samples (p=0.012). Autophagy related genes *LC3A, LC3B, ULK1* and *P62* are enhanced expression in KO cells compared to WT cells, at 0Hrs slight showing slight increase in *LC3A* (p=0.003) and *LC3B* (p=0.005) but *ULK1* (p=0.002) and *P62* (p=0.006) are reduced. At 2 hours showed significant enhanced expression in *LC3A* (p=0.001), *ULK1* (p=0.009) but no significant change observed in *LC3B* (p=0.066) and *P62* (p=0.56). At 4 hours also followed the similar trend *LC3A* (p=0.010), *LC3B* (p=0.002) and *ULK1* (p=0.009) but *P62* (p=0.233) showed no significant difference. At 6 hours all autophagy genes showed greater enhanced expression in KO cells *LC3A* (p<0.001), *LC3B* (p<0.001) and *ULK1* (p<0.001) but *P62 (*p<0.001) significantly reduced. The autophagy regulating gene *mTORC1* followed the similar trend with *CLEC16A* and showing lower expression at 0 hours (p<0.001), 2 hours (p<0.001), 4 hours (p<0.001) and both WT and KO cells showed greater down trend and no significant difference observed (p=0.272). All the measurements were repeated at least three tim s (n=3), calculated mean ± SD, and significance was calculated for pairwise comparisons using GraphPad Prism and multiple t-test per row method.

**Figure 6B.**
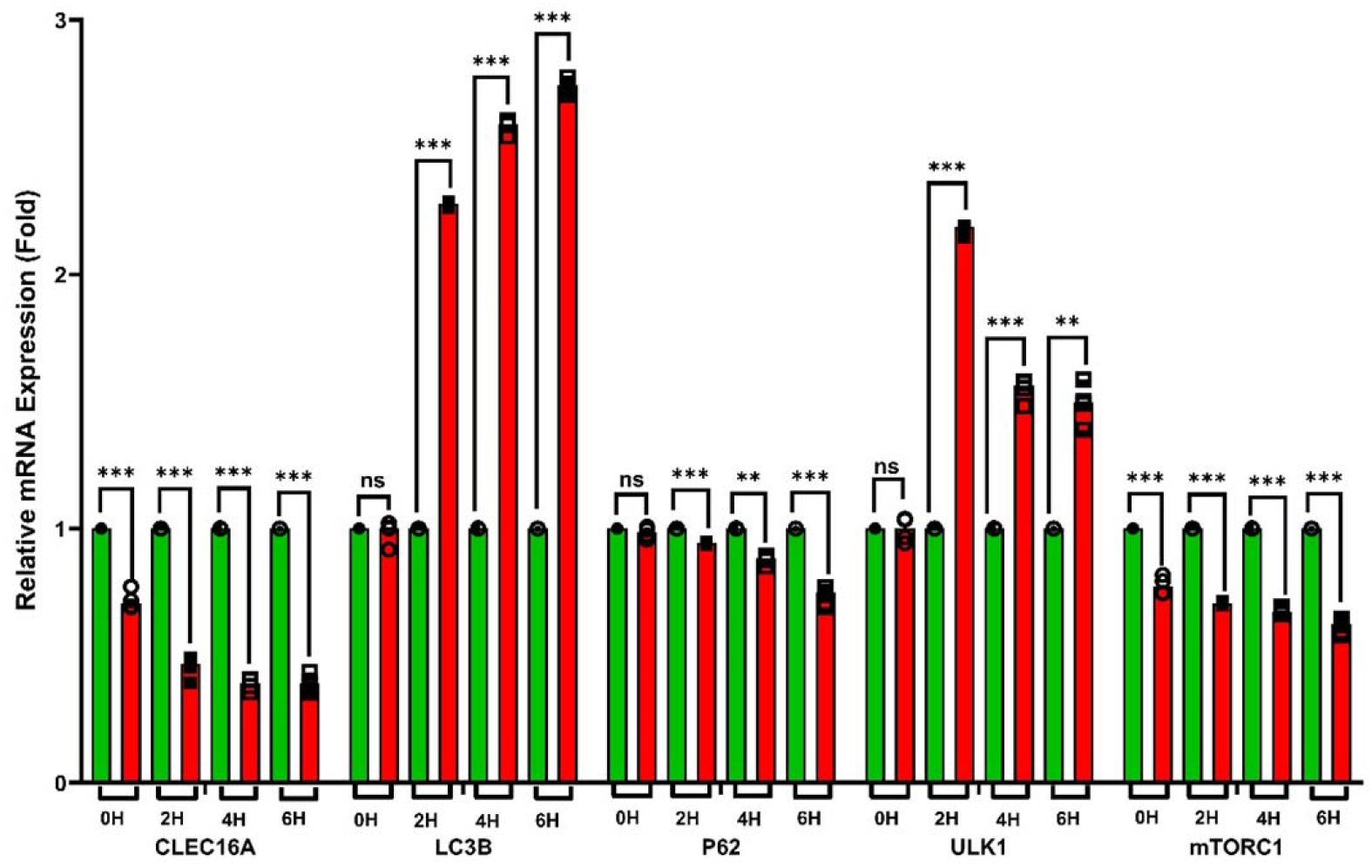
Starvation induced autophagy in CLEC16A knockdown (KD) cells. qRT-PCR assessment of EBSS starved CLEC16A-KD Namalwa cells showing the differential expression of autophagy-related genes. Comparison of EBSS starved CLEC16A-KD cells with WT cells at a regular intervals of 0 Hrs (basal), 2Hrs, 4Hrs and 6Hrs denoting a significant reduction in autophagy-related genes. In KD cells *CLEC16A* gene expression (p<0.001) and *mTORC1* (p<0.001) are reduced but not impacted the other autophagy markers LC3B (p=0.552), P62 (p=0.191), *ULK1* (p=0.866) and compared to wild type WT. Starved KD cells showing significantly enhanced expression of *LC3B* at 2Hr, 4Hr and 6Hr (p<0.001,0.001, 0.001), similarly *ULK1* increased at 2Hr, 4Hr and 6Hr (p<0.001, p<0.001, p<0.01), but *P62* reduced at 2Hr, 4Hr and 6Hr (p<0.001, p<0.01, p<0.001). *CLEC16A* showed significant down-trend at 2Hr, 4Hr and 6Hrs (p<0.001, p<.001, p<0.001) and *mTORC1* also significantly reduced at 2Hr, 4Hr and 6Hr (p<0.001, p<0.001, p<0.001). All the measurements were replicated (n=3) and calculated using multiple t-test method on GraphPad Prism.

At earlier time points (2 and 4 hours), measured transcripts generally showed the same trend but with smaller magnitude of change. Exceptions include *mTORC1*, which showed significant decreases in KO cells at the earlier time points (∼0.35-fold, p<0.0001). Effects of the KO on transcript levels during autophagy were generally consistent with the effects of the KO at baseline (*i*.*e*., 0 hours), but with larger magnitude, suggesting that autophagy progression exacerbates existing differences in transcript levels from the KO. Ex include *ULK1*, which went from a 0.3-fold decrease at baseline to large and significant increases autophagy progression.

Similar trends were observed in CLEC16A knockdown (KD) Namalwa cells compared to WT cells (**Figure 6B**). At baseline, KD cells showed significant reductions in *CLEC16A* (0.3-fold, *p* < 0.0001) and *mTORC1* (0.27- fold, *p* < 0.0001), while no significant changes were observed for other autophagy-related genes (*LC3B, P62*, and *ULK1*). Upon starvation, at 2 hours, *LC3B* and *ULK1* expression increased significantly, while *CLEC16A, P62*, and *mTORC1* were significantly reduced. At 4 and 6 hours, *LC3B* expression continued to increase (1.58- and 1.74-fold, *p* < 0.0001, respectively), and *ULK1* remained elevated (∼0.58-fold, *p* < 0.0001) compared to baseline but slightly decreased compared to the 2-hour time point. Conversely, *CLEC16A* (∼0.61-fold, *p* < 0.0001), *P62* (0.13-fold, *p* = 0.001 at 4 hours; 0.27-fold, *p* < 0.0001 at 6 hours), and *mTORC1* (∼0.35-fold, *p* < 0.0001) exhibited steady and significant declines during prolonged starvation.

As shown in earlier studies, mTORC1 showed lower expression in the nutrient-deprived conditions[23]. Further it was established that CLEC16A – an endolysosomal E3 ubiquitin ligase – plays a pivotal role in mTOR activity[24], consistent with heavy regulation of mTOR by ubiquitin[25]. Together, these findings suggest that CLEC16A enhances mTORC1 activity, in turn resulting in negative regulation of autophagy. These results corroborate an earlier study[24], where elevated expression of CLEC16A contributed to mTORC1 activation and reduced autophagy, while reduced CLEC16A expression delayed mTORC1 activity and enhanced LC3- mediated autophagy.

## Discussion

Here we have identified an intronic SNP in *CLEC16A* with dramatic allele-specific effects on enhancer activity, epigenetic marks, expression of diverse downstream genes, and progression of starvation-induced autophagy, notably through regulation of mTORC1.

The role of CLEC16A (C-type lectin domain family 16, member A – actually an endolysosomal E3 ubiquitin ligase) in cellular processes, particularly autophagy[10,14,15], has been increasingly recognized through diverse studies, including those on its *Drosophila* ortholog, Ema, an endosomal membrane protein essential for endosomal trafficking and maturation[26,27]. The conservation of CLEC16A’s function across species is highlighted by the successful rescue of the *Drosophila Ema* null mutant with human CLEC16A expression (ref). Ema’s critical role in autophagosomal growth and autophagy further underscores CLEC16A’s involvement in these processes.

In mice, Soleimanpour *et al*.[15] and other studies[14,28] have demonstrated that Clec16a deficiency, particularly in mice with pancreas-specific somatic deletions, leads to elevated levels of Parkin—a key regulator of mitophagy—resulting in abnormal mitochondrial morphology and function of pancreatic islets, reduced oxygen consumption, and ATP production. These findings, corroborated in other Clec16a mutant mouse models[29], emphasize CLEC16A’s significant role in autophagy and its broader implications for neurological and metabolic diseases. Thus, CLEC16A is a promising candidate for further functional studies and therapeutic exploration.

The single nucleotide polymorphism (SNP) rs17673553 has emerged as a critical regulatory element influencing gene expression and susceptibility to autoimmune diseases, including SLE[5]. As an expression quantitative trait locus (eQTL) for multiple genes, including *CLEC16A*, rs17673553 modulates enhancer activity, with the risk allele (A) demonstrating higher enhancer activity compared to the non-risk allele (G), resulting in higher levels of CLEC16A transcript and protein. We also showed that rs17673553 directly affects transcription factor binding and histone modifications, impacting gene expression and chromatin interactions.

ChIP-qPCR experiments further reveal that the risk allele is associated with elevated levels of enhancer-associated histone marks, specifically H3K27ac and H3K4me1, which are essential for maintaining an open chromatin state conducive to transcription. The lack of significant differences in H3K4me3, typically associated with promoter regions, suggests that rs17673553 specifically modulates enhancer activity rather than promoter function. Additionally, the differential binding of transcription factors (TFs) such as CTCF, STAT3, and GATA3 to the rs17673553 locus underscores its role in gene regulation and provides mechanistic details. CTCF is involved in various cellular processes, including insulating promoters from enhancers, organizing chromatin structure, regulating long-range chromatin interactions, and maintaining the three-dimensional architecture of the genome. STAT3 and GATA3, heavily involved in immune regulation[30], bind more tightly to the risk A allele, showing how rs17673553 may influence gene expression by altering TF recruitment in a context-dependent manner. Importantly, our results for DNA binding, enhancer activity, and mRNA and protein levels from risk and non-risk genotype cells are consistent, with the risk allele associated with tighter TF binding and levels of transcription and translation.

Our experimental findings are consistent with the notion that CLEC16A negatively regulates starvation-induced autophagy by enhancing mTORC1’s inhibitory effects[24]. In WT cells, starvation led to a moderate increase in autophagy-related gene expression, and a slight upregulation of *CLEC16A* and *mTORC1*. However, KO cells showed a significant upregulation of *LC3A* and *LC3B*, with a corresponding downregulation of *ULK1, P62*, and mTORC1. These results indicate that CLEC16A’s interaction with mTORC1 plays a crucial role in the negative regulation of autophagy, and that the rs17673553 locus is strongly involved. The rs17673553 risk allele, which modulates *CLEC16A* expression, may contribute to dysregulated autophagy, eventually resulting in downstream autoimmune conditions such as SLE.

The association of rs17673553 (16p13) with SLE susceptibility, as identified through multiple genome-wide association studies (GWAS), is supported by our mechanistic insights. The variant influences SLE risk by modulating enhancer function, gene regulation, and protein function, particularly through CLEC16A’s interaction with mTORC1, thus regulating autophagy. The allele-specific modulation of enhancer activity and the impact on autophagy regulation provide a mechanistic explanation for the observed associations between rs17673553 and SLE susceptibility.

In summary, rs17673553 facts as a key regulatory SNP within an active enhancer region, influencing target gene expression through a complex interplay of enhancer activity, histone modifications, and transcription factor binding. Downstream of gene regulation, CLEC16A directly interacts with mTORC1 to regulate autophagy. Elucidating the precise mechanisms by which rs17673553 modulates gene expression could provide valuable insights into SLE and other autoimmune diseases, potentially informing targeted therapeutic strategies. Our findings provide strong evidence that the non-coding intronic variant rs17673553 in *CLEC16A* is functionally significant in SLE susceptibility. The risk allele increases enhancer activity, leading to higher *CLEC16A* expression and potentially affecting additional target genes. This contributes to SLE pathogenesis by impairing immune cell function, disrupting (*i*.*e*., decreasing) autophagy, and eventually promoting autoimmunity.

## Materials and Methods

### Bioinformatics analysis

To identify functional SNPs within the 16p13 locus, we first considered all potential SNPs within the linkage disequilibrium (LD >80%) block that includes rs35032408, a SNP identified in our ImmunoChip-based SLE association study across Asian populations (Korean, Chinese, and Malaysian)[19]. To prioritize functional SNPs in this LD block, we first conducted a comprehensive bioinformatics analysis to identify potential regulatory elements. This analysis utilized publicly available datasets from ENCODE and other regulatory databases to define active enhancers and other regulatory elements. We then employed tools such as RegulomeDB[31] and LDlink[32] to rank these SNPs based on their potential regulatory roles. We also used PredictSNP2 (a consensus classifier combining five prediction tools for variant prioritization: CADD, DANN, FATHMM, FunSeq2, and GWAVA [33].

Our collected data on *cis*-regulatory elements included chromatin accessibility information from ATAC-seq and DNase-I hypersensitivity assays, key histone modifications such as H3K27ac, H3K4me1, and H3K4me3, and binding data for RNA polymerase II (Pol II) and CTCF. We also analyzed transcription factor binding site data from ENCODE to further assess the regulatory potential of each SNP. Subsequently, we utilized expression quantitative trait loci (eQTL) data from GTeX[34] and eQTLgen[35], followed by chromatin conformation (Hi-C) data from ENCODE, to identify likely functional SNPs and their cognate target genes. Additionally, we evaluated the predicted deleteriousness of each SNP using CADD scores, taking a score >13.0 as indicating potential pathogenicity[36].

### Cell culture and transfection

Human cell lines, including Namalwa (B-cell), lymphoblastoid cell lines (LCLs, B-cells), HEK293 (embryonic kidney), and Jurkat (T-cell), were purchased from ATCC and cultured under standard conditions. Cells were periodically screened for correct identity and mycoplasma infection. Cells were transfected with luciferase reporter constructs containing sequences surrounding either the risk or non-risk allele of rs17673553, along with appropriate controls, using Lipofectamine 3000 (Invitrogen) according to the manufacturer’s protocol.

### Luciferase reporter assay

The impact of rs17673553 on enhancer activity was assessed using luciferase reporter assays. Briefly, a 281 bp sequence surrounding rs17673553 was cloned into the pGL4.26 vector (firefly luciferase; Promega). Cells were cultured in growth media prior to transfection at an appropriate confluency. Plasmids were transiently co-transfected alongside pGL4.74 (*Renilla* luciferase; Promega) as a control for transfection efficiency. Cells were lysed 24 hours post-transfection, and luciferase activity was measured using the Dual-Luciferase Reporter Assay System (Promega). Firefly luciferase activity was normalized to *Renilla* luciferase activity to account for transfection efficiency as a proxy for firefly luciferase expression level. All primer sequences and amplicon sequences are shown in **Supplementary Table ST3**.

### CRISPR-activation and inhibition

To regulate target gene expression, CRISPR-activation (CRISPRa) and inhibition (CRISPRi) plasmids were used. We co-transfected the guide RNA-containing plasmid (VectorBuilder) along with CRISPRa plasmids dCas9-VPR and dCas9-TET1 (Addgene), and CRISPRi plasmids dCas9-KRAB-MECP2 and dCas9-DNMT3A (Addgene) in a 1:3 ratio. Samples were harvested 48 hours post-transfection, RNA was extracted using the Quick-RNA mini kit (Zymo Research), and cDNA was synthesized using the iScript (Bio-Rad) system.

### ChIP-qPCR

Coriell cell lines having homozygous risk or non-risk rs17673553 genotype were selected, and chromatin immunoprecipitation (ChIP) assays were performed with the MAGnify (Invitrogen) ChIP kit according to manufacturer instructions. Briefly, cells were washed thoroughly in PBS, incubated in 1% paraformaldehyde for 10 minutes for cross-linking, and quenched by addition of 1.25 mM glycine. Cells were then washed twice in ice-cold PBS, and lysis buffer added along with protease inhibitors provided in the kit. Cell lysates were sonicated using a Covaris e220 sonicator to shear the chromatin, and diluted into appropriate amounts of dilution buffer with protease inhibitors. Diluted chromatin lysate was then incubated with antibodies against H3K27ac, H3K4me1, H3K4me3, RNA polymerase II, CTCF, STAT3 and GATA3. Post-incubation, samples were washed per manufacturer instructions, and cross-linking was reversed in the presence of proteinase K. Eluted DNA was purified using magnetic beads provided in the kit. DNA concentrations of each sample were quantified with quantitative PCR (qPCR.)

### Gene knockdown using siRNA

To knock-down various target genes, siRNA was used. Briefly, 10 µM working concentrations were prepared of siRNAs targeting *STAT3* (sc-29493) and *GATA3* (sc-29331; Santa Cruz Biotechnology) per manufacturer instructions. siRNA constructs were transfected into Namalwa cells using the Neon (Invitrogen) nucleofection system per the manufacturer protocol for Namalwa cells. Samples were collected at 48 hours post-transfection, and RNA was extracted and quantified for target gene expression using qRT-PCR.

### Autophagy assays

Cells were thoroughly washed in PBS and incubated in Earle’s Balanced Salts (EBSS) (Sigma-E2888) to induce starvation and, as a result, autophagy. Samples were collected at regular intervals, namely at 0 hours (basal), 2 hours, 4 hours, and 6 hours. RNA was extracted and quantified for target gene expression using qRT-PCR.

### qPCR

RNA was extracted from transfected cells using the RNeasy Mini Kit (Qiagen). cDNA was synthesized using the SuperScript IV First-Strand Synthesis System (Invitrogen). qPCR was performed using SYBR Green Master Mix (Applied Biosystems) to quantify the expression levels of target genes, including *CLEC16A*. Relative expression levels were calculated using the ΔΔCt method, normalized to 18S RNA.

### Western blotting

Protein lysates were prepared from Namalwa wild-type (WT) and *CLEC16A*-KO (KO) cells using RIPA buffer. Proteins were separated by SDS-PAGE and transferred to PVDF membranes. Membranes were probed with primary antibodies against CLEC16A (sc-398516-H4) or GAPDH (sc-47724; Santa Cruz Biotechnology), followed by horseradish peroxidase (HRP)-conjugated secondary antibodies. Protein bands were visualized using Enhanced Chemiluminescence (ECL) substrate (Bio-Rad) and quantified using ImageJ software.

## Conclusion

Deciphering the mechanistic underpinnings of GWAS loci remains a significant but essential challenge in post-GWAS research to understand disease mechanisms. Our study identifies the non-coding intronic variant rs17673553 in CLEC16A as a critical contributor to SLE susceptibility, primarily through its effects on enhancer function, gene regulation, mTOR interaction, and autophagy modulation. Future research will aim to further elucidate the downstream pathways influenced by this variant and investigate potential therapeutic strategies to counteract its effects. This work underscores the pivotal role of non-coding genetic variants in the complex etiology of autoimmune diseases such as SLE.

## Data Availability

All data produced in the present work are contained in the manuscript

## Disclaimer/Publisher’s Note

The statements, opinions and data contained in all publications are solely those of the individual author(s) and contributor(s) and not of MDPI and/or the editor(s). MDPI and/or the editor(s) disclaim responsibility for any injury to people or property resulting from any ideas, methods, instructions or products referred to in the content.

## References

1. Hoi, A.; Igel, T.; Mok, C.C.; Arnaud, L. Systemic lupus erythematosus. Lancet 2024, 403, 2326–2338, doi:10.1016/s0140-6736(24)00398-2.

2. Akhil, A.; Bansal, R.; Anupam, K.; Tandon, A.; Bhatnagar, A. Systemic lupus erythematosus: latest insight into etiopathogenesis. Rheumatol Int 2023, 43, 1381–1393, doi:10.1007/s00296-023-05346-x.

3. Rodríguez, R.D.; Alarcón-Riquelme, M.E. Exploring the contribution of genetics on the clinical manifestations of systemic lupus erythematosus. Best Pract Res Clin Rheumatol 2024, 101971, doi:10.1016/j.berh.2024.101971.

4. Fazel-Najafabadi, M.; Looger, L.L.; Rallabandi, H.R.; Nath, S.K. A multilayered post-GWAS analysis pipeline defines functional variants and target genes for systemic lupus erythematosus (SLE). Arthritis Rheumatol 2024, doi:10.1002/art.42829.

5. Hakonarson, H.; Grant, S.F.; Bradfield, J.P.; Marchand, L.; Kim, C.E.; Glessner, J.T.; Grabs, R.; Casalunovo, T.; Taback, S.P.; Frackelton, E.C.; et al. A genome-wide association study identifies KIAA0350 as a type 1 diabetes gene. Nature 2007, 448, 591–594, doi:10.1038/nature06010.

6. Zhang, Z.; Cheng, Y.; Zhou, X.; Li, Y.; Gao, J.; Han, J.; Quan, C.; He, S.; Lv, Y.; Hu, D.; et al. Polymorphisms at 16p13 are associated with systemic lupus erythematosus in the Chinese population. J Med Genet 2011, 48, 69–72, doi:10.1136/jmg.2010.077859.

7. Pushkarev, O.; van Mierlo, G.; Kribelbauer, J.F.; Saelens, W.; Gardeux, V.; Deplancke, B. Non-coding variants impact cis-regulatory coordination in a cell type-specific manner. Genome Biol 2024, 25, 190, doi:10.1186/s13059-024-03333-4.

8. Peña-Martínez, E.G.; Rodríguez-Martínez, J.A. Decoding Non-coding Variants: Recent Approaches to Studying Their Role in Gene Regulation and Human Diseases. Front Biosci (Schol Ed) 2024, 16, 4, doi:10.31083/j.fbs1601004.

9. Cano-Gamez, E.; Trynka, G. From GWAS to Function: Using Functional Genomics to Identify the Mechanisms Underlying Complex Diseases. Front Genet 2020, 11, 424, doi:10.3389/fgene.2020.00424.

10. Pearson, G.; Chai, B.; Vozheiko, T.; Liu, X.; Kandarpa, M.; Piper, R.C.; Soleimanpour, S.A. Clec16a, Nrdp1, and USP8 Form a Ubiquitin-Dependent Tripartite Complex That Regulates β-Cell Mitophagy. Diabetes 2017, 67, 265–277, doi:10.2337/db17-0321.

11. Li, J.; Jørgensen, S.F.; Maggadottir, S.M.; Bakay, M.; Warnatz, K.; Glessner, J.; Pandey, R.; Salzer, U.; Schmidt, R.E.; Perez, E.; et al. Association of CLEC16A with human common variable immunodeficiency disorder and role in murine B cells. Nat Commun 2015, 6, 6804, doi:10.1038/ncomms7804.

12. Skinningsrud, B.; Lie, B.A.; Husebye, E.S.; Kvien, T.K.; Førre, Ø.; Flatø, B.; Stormyr, A.; Joner, G.; Njølstad, P.R.; Egeland, T.; et al. A CLEC16A variant confers risk for juvenile idiopathic arthritis and anti-cyclic citrullinated peptide antibody negative rheumatoid arthritis. Ann Rheum Dis 2010, 69, 1471–1474, doi:10.1136/ard.2009.114934.

13. Hoppenbrouwers, I.A.; Aulchenko, Y.S.; Janssens, A.C.; Ramagopalan, S.V.; Broer, L.; Kayser, M.; Ebers, G.C.; Oostra, B.A.; van Duijn, C.M.; Hintzen, R.Q. Replication of CD58 and CLEC16A as genome-wide significant risk genes for multiple sclerosis. J Hum Genet 2009, 54, 676–680, doi:10.1038/jhg.2009.96.

14. Pandey, R.; Bakay, M.; Hakonarson, H. CLEC16A-An Emerging Master Regulator of Autoimmunity and Neurodegeneration. Int J Mol Sci 2023, 24, doi:10.3390/ijms24098224.

15. Soleimanpour, S.A.; Gupta, A.; Bakay, M.; Ferrari, A.M.; Groff, D.N.; Fadista, J.; Spruce, L.A.; Kushner, J.A.; Groop, L.; Seeholzer, S.H.; et al. The diabetes susceptibility gene Clec16a regulates mitophagy. Cell 2014, 157, 1577–1590, doi:10.1016/j.cell.2014.05.016.

16. Liu, X.; Qin, H.; Xu, J. The role of autophagy in the pathogenesis of systemic lupus erythematosus. Int Immunopharmacol 2016, 40, 351–361, doi:10.1016/j.intimp.2016.09.017. Epub 2016 Sep 25.

17. Rockel, J.S.; Kapoor, M. Autophagy: controlling cell fate in rheumatic diseases. Nat Rev Rheumatol 2016, 12, 517–531, doi:10.1038/nrrheum.2016.92.

18. Jin, M.; Zhang, Y. Autophagy and Autoimmune Diseases. Adv Exp Med Biol 2020, 1207, 405–408, doi:10.1007/978-981-15-4272-5_28.

19. Sun, C.; Molineros, J.E.; Looger, L.L.; Zhou, X.J.; Kim, K.; Okada, Y.; Ma, J.; Qi, Y.Y.; Kim-Howard, X.; Motghare, P.; et al. High-density genotyping of immune-related loci identifies new SLE risk variants in individuals with Asian ancestry. Nat Genet 2016, doi:10.1038/ng.3496.

20. Kircher, M.; Witten, D.M.; Jain, P.; O’Roak, B.J.; Cooper, G.M.; Shendure, J. A general framework for estimating the relative pathogenicity of human genetic variants. Nat Genet 2014, 46, 310–315, doi:10.1038/ng.2892. Epub 2014 Feb 2.

21. Yeo, N.C.; Chavez, A.; Lance-Byrne, A.; Chan, Y.; Menn, D.; Milanova, D.; Kuo, C.C.; Guo, X.; Sharma, S.; Tung, A.; et al. An enhanced CRISPR repressor for targeted mammalian gene regulation. Nat Methods 2018, 15, 611–616, doi:10.1038/s41592-018-0048-5. Epub 2018 Jul 16.

22. Konermann, S.; Brigham, M.D.; Trevino, A.E.; Joung, J.; Abudayyeh, O.O.; Barcena, C.; Hsu, P.D.; Habib, N.; Gootenberg, J.S.; Nishimasu, H.; et al. Genome-scale transcriptional activation by an engineered CRISPR-Cas9 complex. Nature 2015, 517, 583–588, doi:10.1038/nature14136.

23. Kim, S.G.; Buel, G.R.; Blenis, J. Nutrient regulation of the mTOR complex 1 signaling pathway. Mol Cells 2013, 35, 463–473, doi:10.1007/s10059-013-0138-2.

24. Tam, R.C.; Li, M.W.; Gao, Y.P.; Pang, Y.T.; Yan, S.; Ge, W.; Lau, C.S.; Chan, V.S. Human CLEC16A regulates autophagy through modulating mTOR activity. Exp Cell Res 2017, 352, 304–312, doi:10.1016/j.yexcr.2017.02.017. Epub 2017 Feb 20.

25. Jiang, Y.; Su, S.; Zhang, Y.; Qian, J.; Liu, P. Control of mTOR signaling by ubiquitin. Oncogene 2019, 38, 3989–4001, doi:10.1038/s41388-019-0713-x.

26. Kim, S.; Wairkar, Y.P.; Daniels, R.W.; DiAntonio, A. The novel endosomal membrane protein Ema interacts with the class C Vps-HOPS complex to promote endosomal maturation. J Cell Biol 2010, 188, 717–734, doi:10.1083/jcb.200911126.

27. Gingerich, M.A.; Liu, X.; Chai, B.; Pearson, G.L.; Vincent, M.P.; Stromer, T.; Zhu, J.; Sidarala, V.; Renberg, A.; Sahu, D.; et al. An intrinsically disordered protein region encoded by the human disease gene CLEC16A regulates mitophagy. Autophagy 2023, 19, 525–543, doi:10.1080/15548627.2022.2080383.

28. Pearson, G.; Chai, B.; Vozheiko, T.; Liu, X.; Kandarpa, M.; Piper, R.C.; Soleimanpour, S.A. Clec16a, Nrdp1, and USP8 Form a Ubiquitin-Dependent Tripartite Complex That Regulates β-Cell Mitophagy. Diabetes 2018, 67, 265–277, doi:10.2337/db17-0321.

29. Pandey, R.; Bakay, M.; Strenkowski, B.P.; Hain, H.S.; Hakonarson, H. JAK/STAT inhibitor therapy partially rescues the lipodystrophic autoimmune phenotype in Clec16a KO mice. Sci Rep 2021, 11, 7372, doi:10.1038/s41598-021-86493-8.

30. Mackie, J.; Ma, C.S.; Tangye, S.G.; Guerin, A. The ups and downs of STAT3 function: too much, too little and human immune dysregulation. Clin Exp Immunol 2023, 212, 107–116, doi:10.1093/cei/uxad007.

31. Dong, S.; Zhao, N.; Spragins, E.; Kagda, M.S.; Li, M.; Assis, P.; Jolanki, O.; Luo, Y.; Cherry, J.M.; Boyle, A.P.; et al. Annotating and prioritizing human non-coding variants with RegulomeDB v.2. Nat Genet 2023, 55, 724–726, doi:10.1038/s41588-023-01365-3.

32. Machiela, M.J.; Chanock, S.J. LDlink: a web-based application for exploring population-specific haplotype structure and linking correlated alleles of possible functional variants. Bioinformatics 2015, 31, 3555–3557, doi:10.1093/bioinformatics/btv402. Epub 2015 Jul 2.

33. Bendl, J.; Musil, M.; Štourač, J.; Zendulka, J.; Damborský, J.; Brezovský, J. PredictSNP2: A Unified Platform for Accurately Evaluating SNP Effects by Exploiting the Different Characteristics of Variants in Distinct Genomic Regions. PLoS computational biology 2016, 12, e1004962, doi:10.1371/journal.pcbi.1004962.

34. The GTEx Consortium atlas of genetic regulatory effects across human tissues. Science 2020, 369, 1318, doi:10.1126/science.aaz1776.

35. Võsa, U.; Claringbould, A.; Westra, H.J.; Bonder, M.J.; Deelen, P.; Zeng, B.; Kirsten, H.; Saha, A.; Kreuzhuber, R.; Yazar, S.; et al. Large-scale cis- and trans-eQTL analyses identify thousands of genetic loci and polygenic scores that regulate blood gene expression. Nat Genet 2021, 53, 1300–1310, doi:10.1038/s41588-021-00913-z.

36. Rentzsch, P.; Witten, D.; Cooper, G.M.; Shendure, J.; Kircher, M. CADD: predicting the deleteriousness of variants throughout the human genome. Nucleic Acids Res 2019, 47, D886–D894, doi:10.1093/nar/gky1016.

